# Multi-center improvement in screening for dystonia in young people with cerebral palsy

**DOI:** 10.1101/2024.09.13.24313431

**Authors:** Bhooma R. Aravamuthan, Emma Lott, Esra Pehlivan, Keerthana Chintalapati, Deborah Grenard, Desiree Roge, Rose Gelineau-Morel, Dante Kyle, Christie Becu, Michael Kruer, Linn Katus, Paul Gross, Amy Bailes, Cerebral Palsy Research Network

**Author notes:** Corresponding Author: Dr. Bhooma R. Aravamuthan, Division of Pediatric Neurology, Department of Neurology, Washington University School of Medicine, 660 South Euclid Avenue, Campus Box 8111 St. Louis MO 63110-1093. **Funding/Support:** NINDS 1K08NS117850-01A1 (BRA). **Role of Funder/Sponsor:** NINDS had no role in the design and conduct of this study.

## Abstract

**Background and Objectives:** Dystonia is a common, debilitating, and often treatment refractory motor symptom of cerebral palsy (CP), affecting 70-80% of this population based on research assessments. However, routine clinical evaluation for dystonia in CP has failed to match these expected numbers. Addressing this diagnostic gap is a medical imperative because the presence of dystonia rules in or out certain treatments for motor symptoms in CP. Therefore, our objective was to optimize rates of clinical dystonia screening to improve rates of clinical dystonia diagnosis.

**Methods:** Using the quality improvement (QI) infrastructure of the Cerebral Palsy Research Network (CPRN), we developed and implemented interventions to increase the documentation percentage of five features of dystonia in young people with CP, aged 3-21 years old. This QI initiative was implemented by seven physiatry and pediatric movement disorders physicians at four tertiary-care pediatric hospitals between 10/10/21 and 7/1/23. We collected visit data cross-sectionally across all participating sites every 2 weeks and tracked our progress using control charts.

**Results:** We assessed 847 unique visits, mostly for established patients (719/847, 85%) who were 9.2 years old on average (95% CI 8.8-9.5). By 4/10/22, the mean percentage of dystonia screening elements documented across all sites rose from 39% to 90% and the mean percentage of visits explicitly documenting the presence or absence of dystonia rose from 65% to 94%. By 10/23/22, the percentage of visits diagnosing dystonia rose from 57% to 74%. These increases were all sustained through the end of the study period in 7/1/23.

**Discussion:** Using a rigorous QI-driven process across four member sites of a North American learning health network (CPRN), we demonstrated that we could increase screening for dystonia and that this was associated with increased clinical dystonia diagnosis, matching expected research-based rates. We propose that similar screening should take place across all sites caring for people with CP.

---

Cerebral palsy (CP) is the most common motor disability across the lifespan affecting 2-4 of every 1000 people.^1^ Dystonia is a common, debilitating, and often treatment refractory movement symptom of CP,^2,3^ affecting 70-80% of this population based on research assessments.^4^ However, routine clinical evaluation for dystonia in CP has failed to match these expected numbers.^5,6^ We have shown that only 13% of children with CP and leg dystonia had that leg dystonia identified in a clinic visit, even when assessed by pediatric movement disorders physicians specializing in CP.^5^

This staggering diagnostic gap is critical to fill for people with CP for many reasons. Dystonia is often associated with significant gross motor functional limitations and pain in people with CP,^2,3^ and requires a distinct treatment regimen.^7^ Dystonia diagnosis is critical to both rule in and rule out certain treatments.^7–9^ For example, the presence of dystonia is a relative contraindication for selective dorsal rhizotomy and can affect the choice of orthopedic procedures in people with CP.^8,9^ Furthermore, early dystonia identification is valuable because targeted surgical treatments like deep brain stimulation have been shown to work best early after dystonia onset.^10^ Therefore, addressing the dystonia diagnostic gap is a medical imperative for people with CP.

Barriers to dystonia identification in the clinic are numerous. Dystonia is variable in its appearance, making it difficult to codify or recognize in a single clinical evaluation.^11–13^ We have shown that dystonia diagnostic rates in people with CP improve with longitudinal evaluation.^5^ This highlights the importance of considering the presence of dystonia at every single clinical visit for a person with CP. The variability of dystonia also highlights the importance of engaging another group of experts who have a more comprehensive and longitudinal view of children with CP: caregivers. We have shown that caregiver descriptions of their child’s movements have high positive predictive value for gold standard dystonia diagnosis.^14^

To facilitate a standardized approach for identifying dystonia in children with CP, we developed and implemented dystonia screening interventions incorporating the expertise of clinicians and caregivers. Using rigorous quality improvement (QI) processes,^15^ we implemented this dystonia screening across four different tertiary care children’s hospitals in the United States, all with dedicated CP and/or movement disorders clinics. Our aims were to determine whether standardized dystonia screening was feasible to incorporate into routine clinical practice and determine whether implementation of this screening would be associated with increased dystonia diagnosis in children with CP.

## Methods

### Cerebral Palsy Research Network Infrastructure

We developed and implemented dystonia screening as the initial initiative of the Cerebral Palsy Research Network (CPRN) Dystonia QI Workgroup. The CPRN is a learning health network of hospitals and community members working together to improve health outcomes for people with CP.^16^ The CPRN hosts both community and clinical registries to gather large volumes of data for research and QI initiatives.

In a concerted effort to prioritize QI, the CPRN held a virtual 2-day QI Bootcamp for all CPRN site investigators participating in QI initiatives within the network. This bootcamp included didactics and interactive scenarios covering topics including construction of Specific, Measurable, Achievable, Relevant, and Time-Bound (SMART) Aims, process mapping, root cause analysis, key driver diagrams, intervention ideation, control charts, and Plan-Do-Study-

Act (PDSA) cycles. Recordings and didactic materials from this Bootcamp remain freely available to all CPRN investigators on our shared Microsoft Teams site (Microsoft Corporation, Redmond, WA).

The CPRN Dystonia QI Workgroup’s Global Aim is to “Optimize care and quality of life for people with CP and dystonia”. To focus this Global Aim to generate our initial SMART aim, we consulted:

1. The CPRN’s published dystonia research priorities for dystonia in CP, which were identified and collectively prioritized by adults with CP, caregivers of people with CP, researchers, and clinicians;^17^ and
2. The CPRN Site Investigators during the virtual April 2021 Annual Investigators Meeting, during which we brainstormed which dystonia research priorities would be most valuable to address first.

Because optimizing care and quality of life for people with CP and dystonia requires first identifying who with CP, in fact, has dystonia, consensus among the CPRN Site Investigators was to first address our published research priority to “improve the clinical consistency of dystonia diagnosis”.^17^

### SMART Aim development

The only clinical dystonia diagnostic aid validated in people with CP (aged 3 years and older) is the Hypertonia Assessment Tool (HAT).^18,19^ The HAT facilitates exam-based differentiation between dystonia and spasticity, the two most common movement symptoms present in people with CP.^20^ Dystonic movements elicited using the HAT can be subtle and can therefore be easy to miss during routine clinical care.^18^ To potentially supplement the HAT for dystonia assessment, we have shown that caregivers’ descriptions of movements could cue clinicians to the presence of dystonia.^14^ Finally, given dystonia’s variable nature, repeated longitudinal assessment for dystonia has been shown to yield higher rates of dystonia diagnosis compared to assessment at a single time point alone.^5^ Taking this data together, we hypothesized we could increase rates of dystonia diagnosis in people with CP by increasing rates of documentation of five dystonia screening elements during all eligible clinic visits:

1. Anywhere in the note: does the patient have dystonia?^5^
2. On history: does the patient/family endorse variable tone?^14^
3. On exam: was there spasticity when checking for velocity-dependent hypertonicity?^19^
4. On exam: was there increased hypertonicity or posturing triggered by voluntary movement?^19^
5. On exam: was there increased hypertonicity or posturing triggered by passive movement, tactile stimulation, or heightened emotion?^19^

Eligible clinic visits were defined as in-person new and follow-up visits where the patient was between 3-21 years old, had an ICD10 visit diagnosis of CP (G80) and was being evaluated in either a CP or movement disorders clinic. The fifth dystonia screening element, which includes dystonia triggers other than voluntary movement, was added in 4/25/22. This element was added after re-affirming that dystonia screening in all children with CP was critical, but not all children with CP consistently have voluntary movements.

Our baseline was assessed across three sites (St. Louis Children’s Hospital, Seattle Children’s Hospital, and Phoenix Children’s Hospital) between 10/10/2021 and 1/1/2022 using up to 2 randomly selected new patient visits and follow-up visits per eligible clinician per site per every 2 week period (as available, noting that clinic schedules vary across sites and clinicians).

Clinicians included four pediatric movement disorders physicians at St. Louis Children’s Hospital, one pediatric physiatrist at Seattle Children’s Hospital, and one pediatric movement disorders physician at Phoenix Children’s Hospital. We additionally spread our QI initiative to one more pediatric movement disorders physician at a fourth site (Children’s Mercy Kansas City) on 9/11/2022. The baseline percentage of documentation of dystonia screening elements 1-4 was 39% (153/388 elements documented across 97 individual clinical visits).

Based on this data, we developed our SMART Aim: Increase the percentage of documented dystonia screening elements in CP or movement disorders clinic visits for people with CP aged 3-21 years old from 39% to 75% between 1/2/22 and 7/02/22.

Our secondary aim was to determine whether the pursuit of our SMART Aim would concomitantly increase the percentage of visits diagnosing dystonia, noting that between 70-80% of children with CP have dystonia based on research assessments.^4^ This secondary aim was developed in 10/23/22 after which data on dystonia diagnosis was routinely captured prospectively and was also retrospectively abstracted for the baseline period (10/10/21 to 1/1/22). Finally, as an additional process measure, we tracked the percentage of visits explicitly documenting the presence or absence of dystonia (dystonia screening element 1). Following the baseline period, we collected data every 2 weeks across all eligible visits performed by clinicians implementing our QI interventions across sites through the end of our study period on 7/1/23.

### Key drivers and interventions

We identified three main key drivers that would affect achievement of our SMART aim and developed multiple interventions to address these drivers (Figure 1):

**Figure 1.**
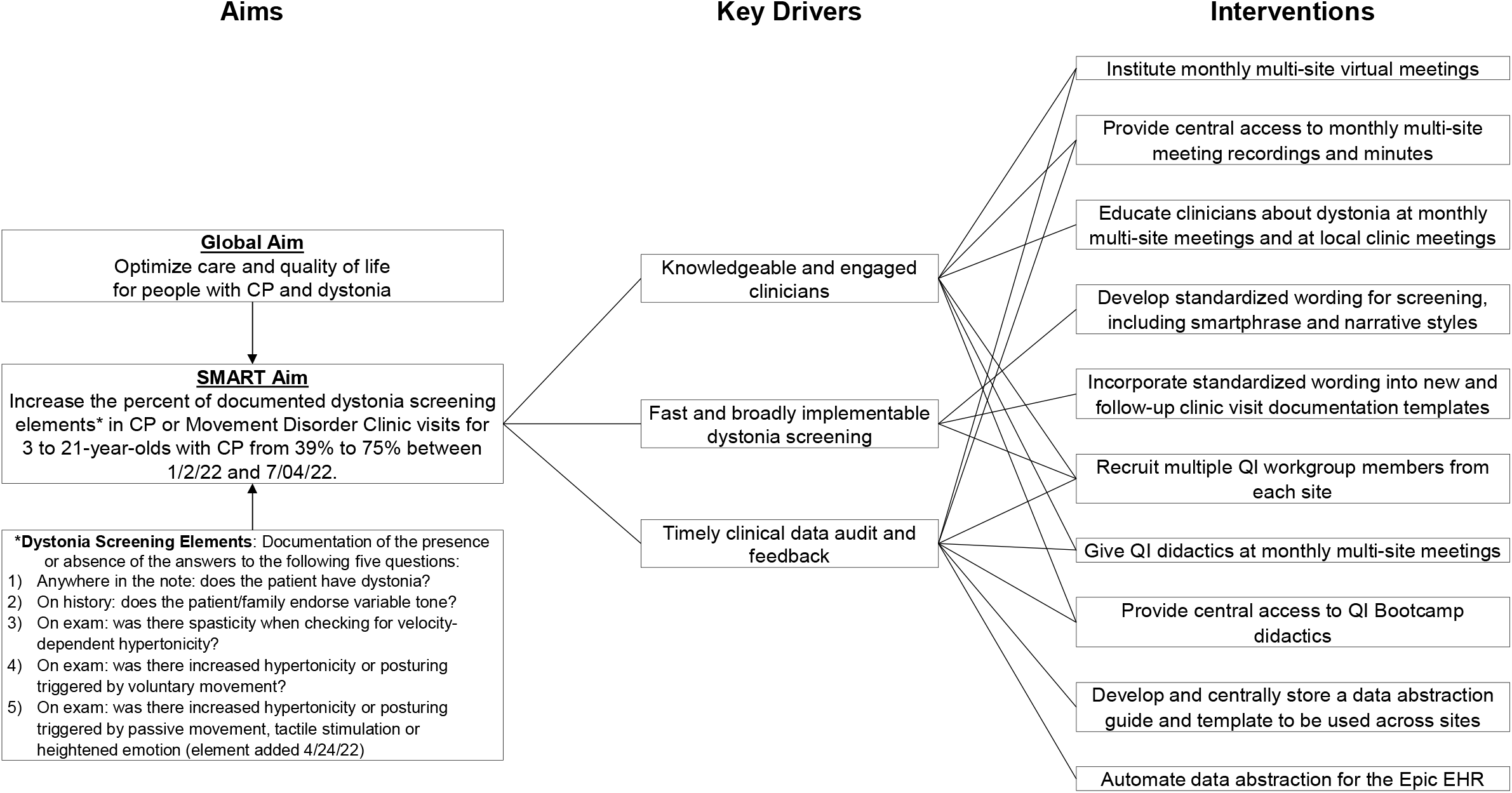
Summary of SMART aim, key drivers, and interventions to optimize dystonia screening. QI – quality improvement; EHR – electronic health record

Key Driver 1: Knowledgeable and engaged clinicians – To ensure a shared clinician knowledge base and promote continued commitment to this QI initiative, we instituted monthly virtual Dystonia QI Workgroup meetings where we would educate clinicians on dystonia and QI related topics (including dystonia underdiagnosis, ^5,6^ the value of longitudinal assessment, ^5^ dystonia clinical features including the value of caregiver expertise, ^14^ and how to develop a SMART aim, key driver diagram, and conduct PDSA cycles). We asked at least one workgroup member from each site to be present at this virtual meetings, but we also recorded these meetings and stored summary minutes on our shared Microsoft Teams site. Workgroup members would then share summaries of dystonia education and QI didactics during their local clinic meetings.

Key Driver 2: Efficient and broadly implementable dystonia screening – Noting limited time in clinic and different documentation styles across clinicians, we provided standardized wording for dystonia screening that could be adapted across sites. Standardized wording prompted documentation of all five dystonia screening elements in both clickable smartphrase and narrative styles (Table 1). We asked clinicians to add this standardized wording to their pre-existing new and follow-up documentation templates. We also recruited multiple QI Workgroup members from each site including trainees and research coordinators so that multiple people at each site knew how to implement standardized wording as a part of documentation.

**Table 1.** Standardized wording to be used in clinical documentation (Intervention 1).

|  | Standardized wording |
| --- | --- |
| **Smartphrase**<br>(example for the Epic electronic health record) | *Cerebral Palsy Research Network Dystonia Assessment*<br>@FNAME@ {DOES/DOES NOT:22499} have dystonia based on my evaluation.<br>•On history, the patient/family {DID/DID NOT:23171} endorse variable tone<br>•On exam, there {WAS/NO:26965} spasticity (when checking for velocity-dependent hypertonicity).<br>•On exam, there {WAS/NO:26965} increased hypertonicity or posturing triggered by voluntary movement.<br>•On exam, there {WAS/NO:26965} increased hypertonicity or posturing triggered by passive movement, tactile stimulation or heightened emotion. |
| **Narrative** | Patient does ***not have dystonia based on my evaluation. On history, they do ***not endorse variable tone. On exam, there was ***not spasticity (when checking for velocity-dependent hypertonicity), and there was ***not increased hypertonicity or posturing triggered by voluntary movement, passive movement, tactile stimulation, or heightened emotion. |

Key Driver 3: Timely clinical data audit and feedback – A large part of the QI-related education given at monthly-site meetings emphasized the importance of timely data audit and feedback to engage in effective PDSA cycles. To further emphasize this, QI Workgroup members were given access to QI Bootcamp recordings that described strategies for timely data audit. To facilitate efficient and standardized data abstraction, we collectively developed a shared data abstraction template and guide and stored this template and anonymized data in our Microsoft Teams site. Finally, we developed a methodology to automate data abstraction from smartlists in the Epic electronic health record (Epic Systems Incorporation, Verona, WI) which facilitated rapid data abstraction at one site.

### Statistical analysis

We tracked our chosen process measures (the percentage of all dystonia elements documented and the percentage of visits explicitly documenting the presence or absence of dystonia) every 2 weeks using control charts templated in Microsoft Excel. The baseline was calculated as the mean across the first 6 data points (10/10/21 to 1/1/22) and recalculated every time there were 8 points above or below the previous mean line (indicating a shift in the data trend).^15^ The upper and lower 3-sigma control limits were calculated as 3 standard deviations above and below each data point.^15,21^

The proportion of visits where a person with CP was documented as having dystonia was compared using Chi-squared tests between the 12-week baseline period and three 12-week periods from 10/23/22 to 7/1/23 following institution of QI interventions (GraphPad Prism, GraphPad Software, Boston, MA). Significance was set *a priori* at p<0.05.

### Standard Protocol Approvals, Registrations, and Patient Consents

This quality improvement study has received human subjects research approval from the Washington University School of Medicine Institutional Review Board (IRB#: 202004133, original approval date 4/28/20). The project is reported in accordance with the Standards for Quality Improvement Reporting Excellence (SQUIRE 2.0). Given that all analyzed data was anonymized and collected during routine clinical care, the IRB granted a waiver of consent.

### Data availability

Anonymized data will be made available by request from any qualified investigator.

## Results

We assessed 847 unique visits across four sites during the study period. These were mostly follow-up visits for established patients (719/847, 85%) aged 9.2 years old on average (95% CI 8.8-9.5). By 4/10/22, the mean percentage of dystonia screening elements documented across all sites rose from 39% to 90% (Figure 2) and the mean percentage of visits explicitly documenting the presence or absence of dystonia rose from 65% to 94% (Figure 3). In the last month of data collection (6/4/23-7/1/23),100% of all dystonia screening elements were documented across all sites. Data points falling below the lower control limits were associated with expanding the documentation ask of clinicians (i.e. adding the fifth dystonia screening element) or spread to other sites or providers (Figure 2). These high percentages were all sustained for over a year through the end of the study period.

**Figure 2.**
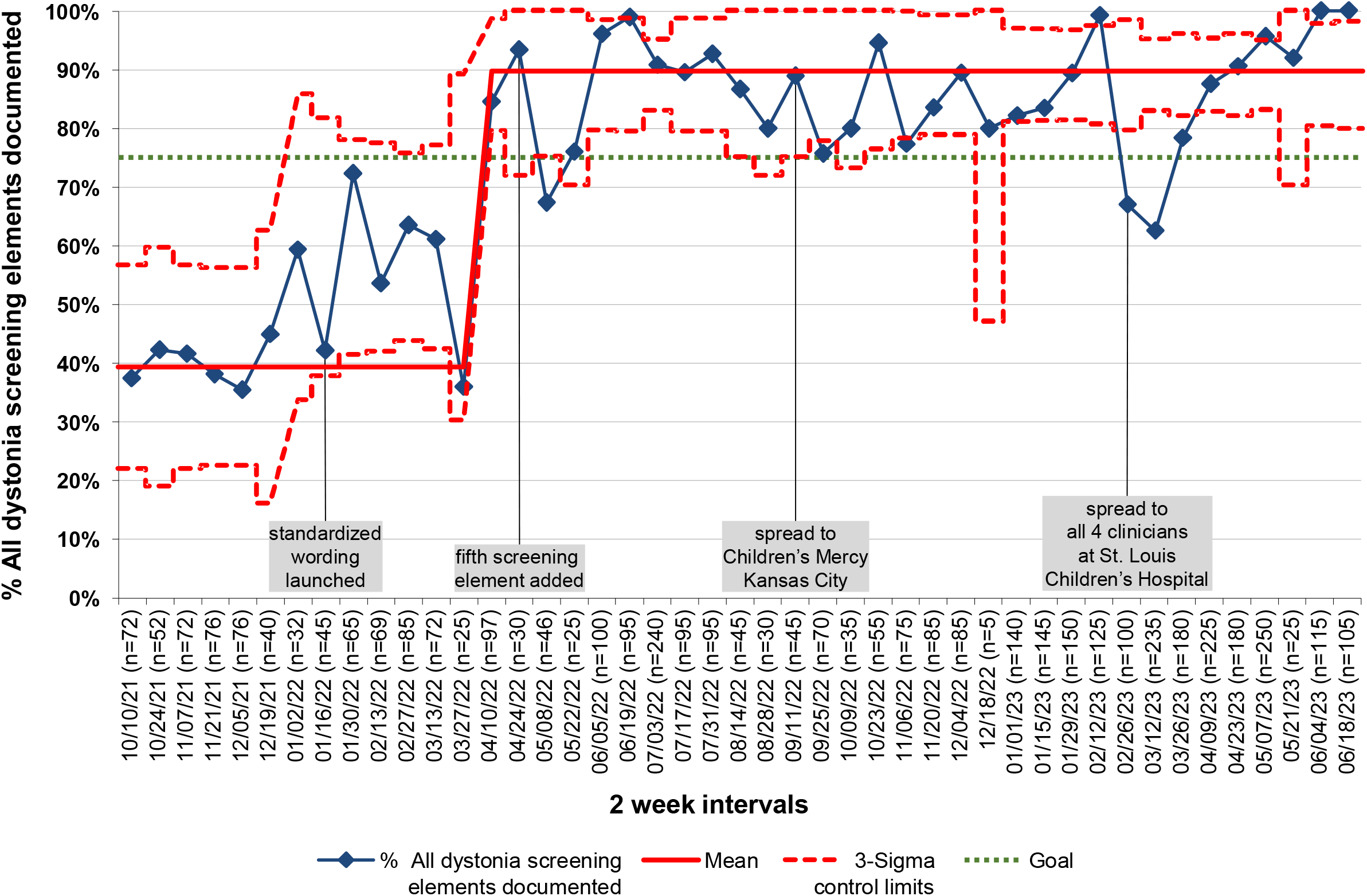
Control chart demonstrating the percentage of all dystonia screening elements documented during the study period.

**Figure 3.**
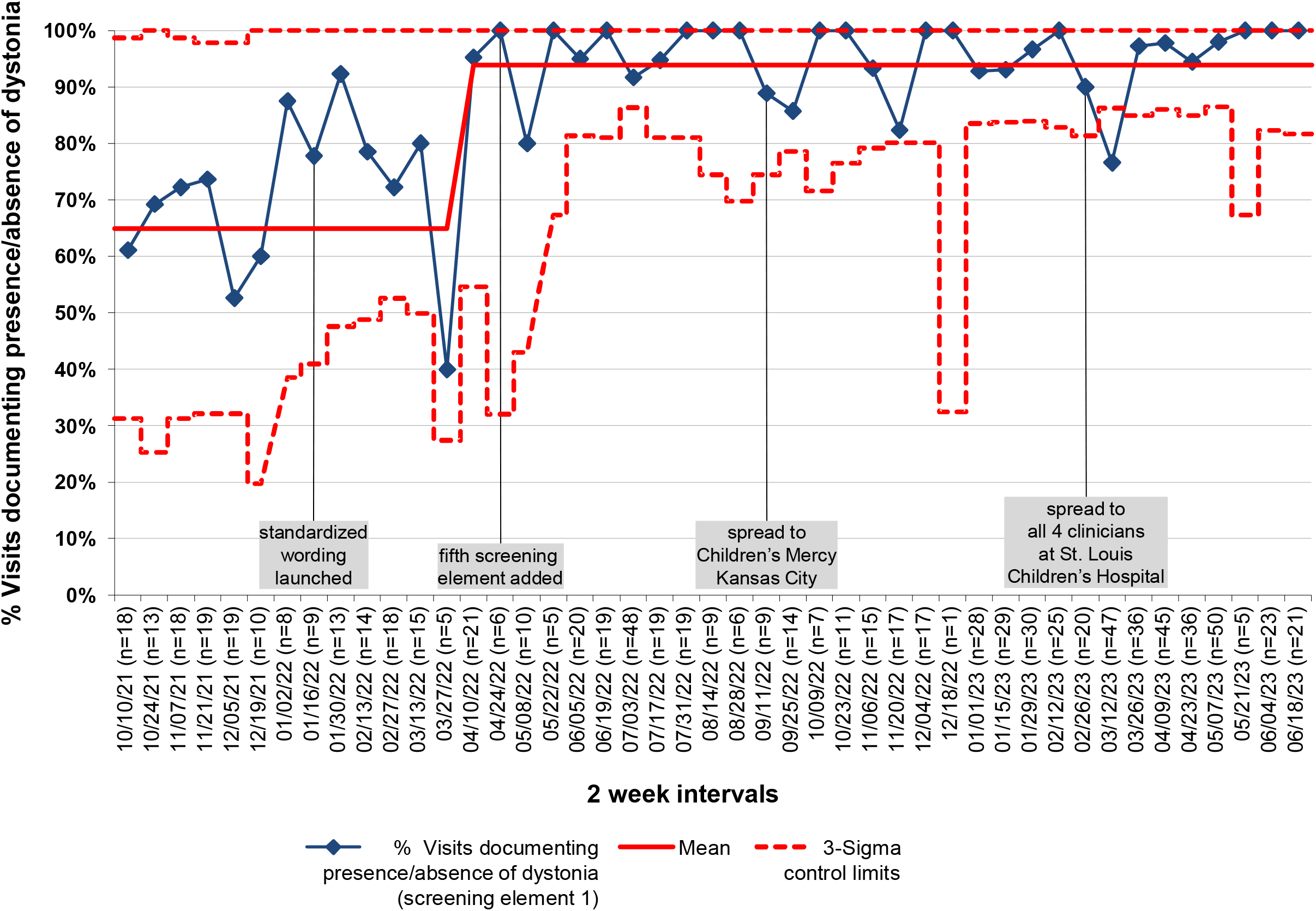
Control chart demonstrating the percentage of visits explicitly documenting the presence or absence of dystonia (dystonia screening element 1) during the study period.

Increased dystonia screening was associated with increased dystonia diagnosis. At baseline, only 57% of all visits documented the presence of dystonia in young people with CP, which is below the expected rate of 70-80%.^4^ By 10/23/22, and sustained through the end of the study period on 7/1/23, the presence of dystonia was documented in 73-76% of all visits (74% on average) which represented a significant increase from baseline (p<0.05, Chi-squared tests). However, when only considering visits that documented the presence or absence of dystonia at all (dystonia screening element 1), the rates of dystonia diagnosis did not change significantly between the baseline (87%) and post-intervention periods (76-82%). What did change significantly was the percentage of visits that explicitly documented the presence or absence of dystonia, which increased from only 65% of visits in the baseline period to between 91-98% in the post-intervention periods assessed (Table 2).

**Table 2.** Visits documenting the presence of dystonia relative to the total number of visits and relative only to the number of visits explicitly documenting the presence or absence of dystonia (dystonia screening element 1, “screening visits”) in young people with CP before and after QI interventions. Superscripts indicate p-values for Chi-squared test comparisons between post-intervention percentages and the corresponding baseline percentages: ***p<0.001, **p<0.005, *p<0.01, ^#^p>0.05, ^##^p>0.2

|  | **Baseline:**<br>10-10-21 to 1-1-22 |  |  | **Post Intervention Period 1:**<br>10-23-22 to 1-14-23 |  |  | **Post Intervention Period 2:**<br>1-15-23 to 4-8-23 |  |  | **Post Intervention Period 3:**<br>4-9-23 to 7-1-23 |  |  |
| --- | --- | --- | --- | --- | --- | --- | --- | --- | --- | --- | --- | --- |
|  | N | % total visits | % screening visits | N | % total visits | % screening visits | N | % total visits | % screening visits | N | % total visits | % screening visits |
| **Yes dystonia** | 55 | 57% | 87% | 68 | 76%* | 82%<sup>##</sup> | 137 | 73%** | 81%<sup>##</sup> | 133 | 74%** | 76%<sup>#</sup> |
| **SCREENING visits** | 63 | 65% | 100% | 83 | 93%*** | 100% | 170 | 91%*** | 100% | 176 | 98%*** | 100% |
| **TOTAL visits** | 97 | 100% | - | 89 | 100% | - | 187 | 100% | - | 180 | 100% | - |

## Discussion

We have demonstrated that documentation of five dystonia screening elements during clinic visits for young people with CP is feasible and is associated with increased dystonia diagnosis. We were able to rapidly achieve documentation of these five screening elements across four sites and were able to collectively sustain this documentation practice for over one year. Our data additionally suggests that increased rates of dystonia diagnosis are more likely to be due to increased rates of screening for dystonia as opposed to increased clinician ability to recognize dystonia in visits where they are already screening for it.

It remains unclear which interventions were most valuable for increasing dystonia diagnosis. It is possible that only some of the five dystonia screening elements we developed were necessary to increase dystonia diagnosis. It is also unclear what role our educational interventions played across sites. These questions can be assessed with future studies examining what subset of our developed interventions are required for sustaining expected rates of dystonia diagnosis in young people with CP.

Future work will address spread to other sites within the CPRN network. Documentation of the five dystonia screening elements is now incorporated into the CPRN registry form and are included in the “highly recommended” registry elements for documentation.^22^ The CPRN is additionally planning network-wide educational initiatives to encourage the documentation of these dystonia screening elements. We can use prospectively collected data from the CPRN registry to assess whether dystonia diagnosis rates increase across the full 35-institution network as the registry forms are updated at each site. This spread will also allow us to determine the utility of these dystonia screening elements in adults.

## Conclusions

Using a rigorous QI-driven process across four member sites of a North American learning health network (CPRN), we demonstrated that we could increase the percentage of CP and movement disorders visits that were screening for dystonia in young people with CP and that this increased screening was associated with increased rates of dystonia diagnosis. Explicitly noting the presence and absence of dystonia has important implications for medical and surgical decision making.^7–9^ Therefore, we propose that similar screening should take place across all sites caring for people with CP.

## Data Availability

All anonymized data produced in the present study are available upon reasonable request to the authors

